# Vaccine effectiveness against SARS-CoV-2 reinfection during periods of Alpha (B.1.1.7), Delta (B.1.617.2) or Omicron (B.1.1.529) dominance: A Danish nationwide study

**DOI:** 10.1101/2022.06.01.22275858

**Authors:** Katrine Finderup Nielsen, Ida Rask Moustsen-Helms, Astrid Blicher Schelde, Mie Agermose Gram, Hanne-Dorthe Emborg, Jens Nielsen, Christian Holm Hansen, Michael Asger Andersen, Marianna Meaidi, Jan Wohlfahrt, Palle Valentiner-Branth

## Abstract

**Introduction:** Individuals with a prior severe acute respiratory corona virus 2 (SARS-CoV-2) infection have a moderate to high degree of protection against reinfection, though seemingly less so when the Omicron variant of SARS-CoV-2 started to circulate. The aim of this study was to evaluate the vaccine effectiveness (VE) against SARS-CoV-2 reinfection, that is, in individuals with prior SARS-CoV-2 infection, during periods with different dominant SARS-CoV-2 variants.

**Methods:** A nationwide cohort study design including all individuals with a confirmed SARS-CoV-2 infection, who were alive and residing in Denmark between 1 January 2020 and 31 January 2022 were used. Using Danish nationwide registries, we obtained information on SARS-CoV-2 infections, Coronavirus Disease 2019 (COVID-19) vaccination, age, sex, comorbidity, staying at hospital and region of affiliation. The study population included were individuals with prior SARS-CoV-2 infection. Crude and adjusted estimates of VE against SARS-CoV-2 reinfection with 95% confidence intervals (CIs) were calculated using Poisson and Cox regression models, respectively. The VE estimates were calculated separately for three periods with different dominant SARS-CoV-2 variants (Alpha (B.1.1.7), Delta (B.1.617.2), or Omicron (B.1.1.529)) and by time since vaccination using unvaccinated as the reference.

**Findings:** The study population comprised of 209,814 individuals infected before or during the Alpha period, 292,978 before or during the Delta period and 245,530 before or during the Omicron period. Of these, 40,281 individuals had completed their primary vaccination series during the Alpha period (19.2%), 190,026 during the Delta period (64.9%) and 158,563 during the Omicron period (64.6%). VE against reinfection following any COVID-19 vaccine type administered in Denmark, peaked at 85% (95% CI: 37% to 97%) at 104 days or more after vaccination during the Alpha period, 88% (95% CI: 81% to 92%) 14-43 days after vaccination during the Delta period and 60% (95% CI: 58% to 62%) 14-43 days after vaccination during the Omicron period. Waning immunity was observed, and was most pronounced during the Omicron period.

**Interpretation:** This study shows that, in previously infected individuals, completing a primary vaccination series was associated with a significant protection against SARS-CoV-2 reinfection compared with no vaccination for all three variant periods. Even though vaccination seems to protect to a lesser degree against reinfection with the Omicron variant, these findings are of public health relevance as they show that previously infected individuals still benefit from COVID-19 vaccination in all three variant periods.

## Introduction

Previous observational studies have investigated the association between Coronavirus Disease 2019 (COVID-19) vaccination and severe acute respiratory corona virus 2 (SARS-CoV-2) reinfection,^1–3^ but the duration and effect of protection from vaccination after a SARS-CoV-2 infection remains uncertain. Despite an estimated moderate to high natural protection against reinfection with non-Omicron variants of SARS-CoV-2,^4–6^ data from Denmark and Qatar suggests a lower protection against reinfection with the Omicron (B.1.1.529) variant.^5,6^ Therefore, it is of great public health concern to examine the additional benefits of vaccination among individuals with a history of SARS-CoV-2 infection.

In Denmark, the healthcare system provides universal healthcare to everyone residing in Denmark,^7^ guaranteeing access to free COVID-19 testing and vaccines as well as medical care. A COVID-19 vaccination program was rolled out in increments from end of December 2020, prioritizing those with increased exposure to SARS-CoV-2 or risk of severe COVID-19.^8^ A booster vaccination campaign was rolled out in the same manner from September 2021. Vaccines administered in Denmark were: Comirnaty (BNT162b2), Spikevax (mRNA-1273), Vaxzevria (ChAdOx1) and Janssen (Ad26.COV2-S).

The study objective was to examine VE against SARS-CoV-2 reinfection in previously infected individuals, and to assess the effect of time since vaccination (waning) in calendar periods where the SARS-CoV-2 variants Alpha (B.1.1.7), Delta (B.1.617.2) or Omicron were dominant.

## Methods

### Data extraction and preparation

The Danish Civil Registration System (CRS) holds information on date of birth, emigration, immigration and death of all individuals in Denmark.^9^ CRS also holds a unique personal registration number for all residents in Denmark. Information on SARS-CoV-2 infections, defined as a positive SARS-CoV-2 Reverse Transcription Polymerase Chain Reaction (RT-PCR) test, was obtained from the Danish Microbiology Database (MiBa), which is a national database containing real-time information on all microbiological laboratory test results from all clinical microbiology and private test centers, including date of sampling.^10^ Information on all COVID-19 vaccines (exposure) was obtained from the Danish Vaccination Registry (DVR). All vaccinators are obliged to document administered vaccines in this registry.^11^ Information on comorbidity was retrieved from the Danish National Patient Registry (DNPR).^12^ A primary vaccination series was defined as two doses COVID-19 mRNA vaccine (Comirnaty or Spikevax), two doses Vaxzevria or one dose Janssen. We combined information from the CRS, DVR and DNPR using the unique personal registration number and identified all individuals who were alive and residing in Denmark between 1 January 2020 and 31 January 2022. Using the unique personal registration number and MiBa, we found those who had a confirmed SARS-CoV-2 infection during the study period, and these individuals constitute the study population. Rapid antigen test results are also recorded in MiBa, but were not included in the analyses due to a low sensitivity.^13^ Individuals with a positive antigen test were urged to confirm the result by RT-PCR. We applied a 90-day window following a laboratory confirmed RT-PCR SARS-CoV-2 positive test to avoid ongoing infections being misclassified as new infections.

The potential confounders age, sex, comorbidity, region of affiliation and staying at hospital were included in the analyses. Information on age, sex, and region of affiliation was obtained from the CRS.^9^ Comorbidity was defines as having a comorbidity diagnosis within 5 years prior to study entry. Diagnoses in the DNPR are coded according to the International Classification of Diseases, 10^th^ revision (ICD- 10).^12^ The ICD-10 codes used to define comorbidity diagnoses are shown in Supplementary Table 1. Categorization of region of affiliation is shown in Supplementary Table 2.

### Statistical analyses

Analyses were performed in three calendar periods with different dominant SARS-CoV-2 variants. A variant period (Alpha, Delta or Omicron) was defined when a variant accounted for 75% or more of all whole genome sequenced PCR tests.^14^ Individuals with a confirmed first-time SARS-CoV-2 infection were followed from 20 February 2021 until 15 June 2021 for the Alpha period, from 4 July to 20 November 2021 for the Delta period, and from 21 December 2021 to 31 January 2022 for the Omicron period. For all three variant periods, individuals were followed from start of follow-up or the date of the first infection, whichever came latest, and until they had a confirmed SARS-CoV-2 reinfection (outcome), died, immigrated, received a booster vaccine or end of follow-up, whichever came first. Separate analyses were conducted for each variant period.

We used a Poisson regression model accounting for over-dispersion (quasi-Poisson) to estimate crude incidence rate ratios (IRRs), and a Cox proportional hazards regression model with underlying calendar time to estimate hazard ratios (HRs) adjusted for sex, age, region of affiliation, any comorbidity and staying at hospital. Sex, any comorbidity, and region of affiliation were included as categorical variables, while age and staying at hospital were included as time-varying covariates. The explanatory variable vaccination was included as a time-varying exposure, and a person was considered completely vaccinated from 14 days or more after the last dose of a primary vaccination series, while a person was unvaccinated until receiving the first vaccine dose. The time from receiving the first vaccine dose and until 13 days after receiving the second dose was excluded from the analyses. We found the protection from previous infection to be stable through the study period (data not shown), and therefore did not include time since infection in the models.

The proportional hazards assumptions were assessed graphically and found to be valid. VE was calculated as a percentage *VE*_*crude*_ = (1 − *IRR*). 100, and *VE*_*adjusted*_ = (1 − *HR*). 100.

Data were analyzed using R version 4.1.2 (R Foundation for Statistical Computing, https://www.R-project.org/).

### Ethical considerations

In Denmark, approval from the Ethics Committee is not required for this type of study. The study adheres to the Strengthening the Reporting of Observational Studies in Epidemiology (STROBE).^15^

## Results

### Study population

The included populations for the variant periods comprised of 209,814 individuals with a COVID-19 infection before or during the Alpha period, 292,978 before or during the Delta period, and 245,530 before or during the Omicron period. Of these individuals, 40,281 (Alpha, 19.2%), 190,026 (Delta, 64.9%) and 158,563 (Omicron, 64.6%) completed their primary vaccination series during follow-up in the respective variant periods (Table 1). The mRNA vaccines, Comirnaty and Spikevax, accounted for more than 84% of the COVID-19 vaccines administered (Table 1). Individuals who completed a primary vaccination series in the Alpha period were older, predominantly female and more individuals had comorbidity when compared to the Delta and Omicron periods. Individuals completing a primary vaccination series in the Omicron period had the lowest median age (Table 1).

**Table 1.** Descriptive overview of the study population at start of follow-up.

| Variant: | Alpha<br>(B.1.1.7) |  | Delta<br>(B.1.617.2) |  | Omicron<br>(B.1.1.529) |  |
| --- | --- | --- | --- | --- | --- | --- |
| Follow-up: | 20 February – 15 June 2021 |  | 4 July – 20 November 2021 |  | 21 December 2021 – 31 January 2022 |  |
|  | Study population at entry (individuals with prior first-time SARS-CoV-2 infection) | Vaccinated individuals | Study population at entry (individuals with prior first-time SARS-CoV-2 infection) | Vaccinated individuals | Study population at entry (individuals with prior first-time SARS-CoV-2 infection) | Vaccinated individuals |
| **Number of individuals included, n (%)** | 209,814 (100) | 40,281 (19.2) | 292,978 (100) | 190,026 (64.9) | 245,530 (100) | 158,563 (64.6) |
| **Sex, n** |  |  |  |  |  |  |
| Female (%) | 106,717 (50.9) | 24,200 (60.1) | 146,111 (49.9) | 95,623 (50.3) | 120,432 (49.0) | 77,595 (48.9) |
| Male | 103,097 | 16,081 | 146,867 | 94,403 | 125,098 | 80,968 |
| **Median age [IQR]** | 35.3 [21.1-52.9] | 63.5 [45.7-73.6] | 32.0 [20.0-50.0] | 41.2 [25.2-55.4] | 25.4 [15.4-38.1] | 27.6 [18.8-40.8] |
| **Vaccine product, n (%)** |  |  |  |  |  |  |
| Comirnaty |  | 34,383 (85.4) |  | 161,115 (84.8) |  | 134,046 (84.5) |
| Janssen |  | 1,057 (2.6) |  | 1,629 (0.9) |  | 466 (0.3) |
| Spikevax |  | 4,781 (11.9) |  | 27,201 (14.3) |  | 24,020 (15.1) |
| Vaxzevria |  | 60 (0.1) |  | 81 (0.04) |  | 31 (0.02) |
| **Region of affiliation, n (%)** |  |  |  |  |  |  |
| Denmark | 150,622 (71.8) | 34,209 (85.0) | 205,051 (70.0) | 147,281 (77.5) | 159,224 (64.8) | 115,417 (72.8) |
| High-income | 45,237 (21.6) | 4,087 (10.1) | 67,875 (23.2) | 31,524 (16.6) | 67,252 (27.4) | 32,932 (20.8) |
| Other | 13,670 (6.5) | 9 (0.02) | 19,896 (6.8) | 11,182 (5.9) | 18,963 (7.7) | 10,179 (6.4) |
| Unknown | 285 (0.1) | 1,976 (5.0) | 156 (0.05) | 39 (0.02) | 91 (0.04) | 35 (0.2) |
| **Any comorbidity, n (%)** | 66,318 (31.6) | 22,360 (55.5) | 85,200 (29.1) | 60,571 (31.9) | 56,710 (23.1) | 35,524 (22.4) |

Abbreviations: IQR, interquartile range; SARS-CoV-2, severe acute respiratory syndrome coronavirus 2

### SARS-CoV-2 reinfection

During the Alpha period, 437 individuals had a confirmed SARS-CoV-2 reinfection (Table 2). The adjusted VE against reinfection was statistically insignificant (42%, 95% CI: -2% to 67%) 14-43 days after vaccination, but rose to significant levels of 84% (95% CI: 65% to 93%) 44-103 days after vaccination and 85% (95% CI: 37% to 97%) 104 days or more after vaccination (Table 2). During the Delta period, 1,678 individuals had a reinfection, giving an adjusted VE against reinfection of 88% (95% CI: 81% to 92%) 14-43 days after vaccination. VE declined from the initial 88% to 59% (95% CI: 41% to 71%) at 134-163 days after vaccination, after which it fluctuated due to few events and was statistically insignificant at 284 days or more after vaccination (Table 2). For infection with the Omicron variant, VE peaked at 60% (95% CI: 58% to 62%) 14-43 days after vaccination and declined to 14% (95% CI: 10% to 17%) 164-193 days after vaccination (Table 2). Here, the VE fluctuated due to few events and became statistically insignificant at 284-313 days after vaccination. VE decreased over time since vaccination for the Delta and Omicron and waning seemed to be more pronounced for the latter.

**Table 2.** Vaccine effectiveness against SARS-CoV-2 reinfection during periods of Alpha, Delta or Omicron dominance.

|  | Alpha variant<br>(B.1.1.7) |  |  |  |  |
| --- | --- | --- | --- | --- | --- |
|  | 20 February - 15 June 2021 |  |  |  |  |
|  | Reinfections, n | PYRS | Adjusted vaccine effectiveness against SARS-COV-2 reinfection* |  |  |
|  |  |  | VE | 95% CI |  |
| Previously infected and unvaccinated | 405 | 41,774.00 | 0% (ref) | - | - |
| Previously infected and vaccinated<br>(Days since vaccination) |  |  |  |  |  |
| 14-43 | 22 | 2,098.77 | 42% | -2% | 67% |
| 44-103 | 8 | 1,778.44 | 84% | 65% | 93% |
| ≥104 | 2 | 406.34 | 85% | 37% | 97% |

Abbreviations: CI, confidence interval; PYRS, person-years; SARS-CoV-2, severe acute respiratory syndrome coronavirus 2; VE, vaccine effectiveness \*Adjusted for age, sex, region of affiliation, any comorbidity and staying at hospital

|  | Delta variant<br>(B.1.617.2) |  |  |  |  |
| --- | --- | --- | --- | --- | --- |
|  | 4 July - 20 November 2021 |  |  |  |  |
|  | Reinfections, n | PYRS | Adjusted vaccine effectiveness against SARS-COV-2 reinfection* |  |  |
|  |  |  | VE | 95% CI |  |
| Previously infected and unvaccinated | 1392 | 33,620.05 | 0% (ref) | - | - |
| Previously infected and vaccinated<br>(Days since vaccination) |  |  |  |  |  |
| 14-43 | 26 | 12,263.52 | 88% | 81% | 92% |
| 44-73 | 32 | 12,918.54 | 85% | 79% | 90% |
| 74-103 | 69 | 11,813.05 | 79% | 72% | 84% |
| 104-133 | 67 | 7,556.37 | 71% | 62% | 78% |
| 134-163 | 44 | 3,796.79 | 59% | 41% | 71% |
| 164-193 | 23 | 2,265.85 | 72% | 54% | 83% |
| 194-223 | 15 | 1,046.61 | 67% | 42% | 81% |
| 224-253 | 5 | 477.28 | 71% | 26% | 88% |
| 254-283 | 3 | 208.60 | 82% | 40% | 94% |
| ≥284 | 2 | 25.22 | 54% | -93% | 89% |

Abbreviations: CI, confidence interval; PYRS, person-years; SARS-CoV-2, severe acute respiratory syndrome coronavirus 2; VE, vaccine effectiveness \*Adjusted for age, sex, region of affiliation, any comorbidity and staying at hospital

**Table 2. Continued**
| **Omicron variant**<br>**(B.1.1.529)** |  |  |  |  |  |
| --- | --- | --- | --- | --- | --- |
| 21 December 2021 - 31 January 2022 (end of follow-up) |  |  |  |  |  |
|  | Reinfections | PYRS | Adjusted vaccine effectiveness against<br>SARS-COV-2 reinfection* |  |  |
|  |  |  | VE | 95% CI |  |
| Previously infected and unvaccinated | 24,002 | 7,712.27 | 0% (ref) | - | - |
| Previously infected and vaccinated<br>(Days since vaccination) |  |  |  |  |  |
| 14-43 | 2033 | 1,690.69 | 60% | 58% | 62% |
| 44-73 | 1271 | 746.05 | 50% | 47% | 53% |
| 74-103 | 1061 | 702.72 | 43% | 39% | 46% |
| 104-133 | 3014 | 1,954.08 | 34% | 32% | 37% |
| 134-163 | 6737 | 3,271.95 | 20% | 17% | 22% |
| 164-193 | 3195 | 1,189.79 | 14% | 10% | 17% |
| 194-223 | 502 | 209.88 | 21% | 13% | 28% |
| 224-253 | 161 | 71.18 | 22% | 8% | 35% |
| 254-283 | 53 | 32.57 | 41% | 22% | 55% |
| 284-313 | 32 | 17.06 | 29% | -1% | 50% |
| 314-343 | 25 | 13.09 | 28% | -8% | 52% |
| ≥344 | 20 | 6.16 | 32% | -7% | 56% |

Abbreviations: CI, confidence interval; PYRS, person-years; SARS-CoV-2, severe acute respiratory syndrome coronavirus 2; VE, vaccine effectiveness \*Adjusted for age, sex, region of affiliation, any comorbidity and staying at hospital

VE against hospitalization and death were also analyzed, but due to too few events, it was not possible to estimate VE.

## Discussion

In this nationwide, population-based cohort study, we found primary COVID-19 vaccination to be associated with a significant VE against SARS-CoV-2 reinfection during periods dominated by Alpha, Delta or Omicron variants.

For the Delta and Omicron periods, the protection was highest 14-43 days after completed primary vaccination series, although lower against the Omicron variant. In the Alpha dominated period, the VE was initially low and statistically insignificant, but rose 104 days or more after vaccination. During this period the oldest and most vulnerable individuals were vaccinated, including those with a less responsive adaptive immune system and antibody production.^16–18^ A slower immune response following vaccination might explain the statistically insignificant VE seen in the immediate post vaccination period in the present study. In addition, a slower clearance of previous infections have been observed the elderly Danish population^4^. Hence, a minimum of 90 days between two positive tests might not be sufficient to clear a SARS-CoV-2 infection in the elderly, which would lead to an underestimated VE estimate in this population.

In the Delta period, a VE of 88% against reinfection was observed 14-43 days after vaccination. This is in accordance with observational studies from Sweden and Israel, in periods of both Alpha and Delta^19^ or Delta dominance^2^ where VE ranged from 66% (95% CI: 61% to 69%)^19^ to 82% (95% CI: 80% to 84%)^2^.

For the Omicron period, our study showed an initial VE against reinfection of 60% (95% CI: 58% to 62%), which is lower than what we found for the other variants, but still indicates an additional protection following vaccination. This is similar to a pre-print study from Qatar, were a VE of 55.1% (95% CI: 50.9% to 58.9%) against reinfection with the Omicron variant after two doses of Comirnaty was estimated.^20^ A lower VE of two doses COVID-19 mRNA vaccines against hospitalization was also seen in a period dominated by Omicron (34.6%, 95% CI: 25.5% to 42.5%) compared to Delta (47.5%, 95% CI: 38.8% to 54.9%), in a test-negative design study from USA.^21^

A major strength of this study is the completeness of the Danish registries, which reduces the risk of selection bias as they cover all individuals residing in Denmark and their contacts with vaccination- or test centers, as well as the Danish personal registration number ensuring individual-level linkage of data. Also, Denmark has had one of the highest testing rates in the world,^22^ which limits the risk of undiscovered reinfections.

This study also has some limitations. Despite the high test rate, we cannot rule out undetected reinfections, especially asymptomatic infections among vaccinated individuals, which might inflate the VE. We also cannot rule out that vaccinated and unvaccinated individuals had different health seeking behavior or risk behavior, which could affect VE. Additionally, from April 2021, a corona pass was introduced in Denmark and a valid corona pass gained by previous infection or vaccination, was required for a broad range of social activities, including restaurants, gyms etc.. This may have affected test activity differently for unvaccinated and vaccinated individuals. However, this was common practice during both the Delta and Omicron period and might therefore not play a role when considering VE in these periods.

In summary, this study showed that among previously infected individuals who have completed a primary vaccination series, there is a significant VE against SARS-CoV-2 reinfection for the SARS-CoV- 2 variants Alpha, Delta and Omicron; lasting up to 9 months. Even though vaccination seems to protect to a lesser degree against reinfection with the Omicron variant, these findings are of public health relevance as they show that previously infected individuals still benefit from COVID-19 vaccination in all three variant periods.

## Data Availability

The data included in this research is part of the Danish national COVID-19 surveillance system database at Statens Serum Institut. The data are available for research upon reasonable request and with permission from the Danish Data Protection Agency and Danish Health and Medicines Authority.

## Supplementary

**Table S1.** Overview of ICD-10 codes included in the comorbidity variable.

| Disease | ICD-10 codes |
| --- | --- |
| Diabetes | E10-E14 |
| Adiposity | E65-E68 |
| Cancer | C00-C80, Z85 |
| Neurological diseases | G10-G14, G20-G23, G35-G37, G71-G73, G80-G83, G90-G91, G93-G96, G99, M51 (without G360, G902) |
| Kidney diseases | N180-N200Z, N18, Z992 |
| Hematological cancers | C81-C96, Z856-Z857 |
| Cardiovascular diseases | I20, I230-I259Z, I45-I499Z, I24, I50-I099Z, I340-I399Z, I05, I44, I21-I238Z, I000-I029Z, I30-I399Z, I26-I289Z, I40-I439Z, I50-I528Z, R01-R012B, I10-I59Z |
| Respiratory diseases | J400-J998Z, J40, J100-J229Z, J68, J430-J499 |
| Immune diseases | B200-B249Z, B20, Z21, D800-D899Z, Z923, Z926, Z941-Z949, Z94 (without Z945, Z947) |
| Other diseases | K700-K709, K70, E150-E909Z, D500-D649Z, D709-D779Z, D50, D650-D699Z, K710-K778Z, Q200-Q349Z, A150-A199Z, A15, Z902, Z905, Y90, Y900-Y919, Z251, |

Abbreviations: ICD-10, International Classification of Diseases 10th revision

**Table S2.** Categorization of region of affiliation.

| Region of affiliation | Definition |
| --- | --- |
| Danish | Individuals who were born in Denmark or abroad and have at least one parent who is Danish citizen and born in Denmark. |
| High-income | Individuals with country of origin* of Nordic countries, EU countries, Andorra, Liechtenstein, Monaco, San Marino, Switzerland, the United Kingdom, the Vatican City, Canada, the United States, Australia and New Zealand. |
| Other | Individuals with country of origin* of all other countries than the countries defined by western heritage. |

\*Country of origin is defined as follows:

- When neither parent is known, the country of origin is defined on the basis of the person’s own information. If the person is an immigrant, it is assumed that the country of origin is equal to the country of birth. If the person is a descendant, it is assumed that the country of origin is equal to the country of citizenship.
- When only one parent is known, the country of origin is defined based on its country of birth. If this is Denmark, the country of citizenship is used.
- When both parents are known, the country of origin is defined on the basis of the mother’s country of birth and country of citizenship, respectively.

## References

1. Gazit S, Shlezinger R, Perez G, et al. The Incidence of SARS-CoV-2 Reinfection in Persons With Naturally Acquired Immunity With and Without Subsequent Receipt of a Single Dose of BNT162b2 Vaccine : A Retrospective Cohort Study. Ann Intern Med. Published online February 15, 2022. doi:10.7326/M21-4130

2. Hammerman A, Sergienko R, Friger M, et al. Effectiveness of the BNT162b2 Vaccine after Recovery from Covid-19. N Engl J Med. Published online February 16, 2022:NEJMoa2119497. doi:10.1056/NEJMoa2119497

3. Cavanaugh AM, Spicer KB, Thoroughman D, Glick C, Winter K. Reduced Risk of Reinfection with SARS-CoV-2 After COVID-19 Vaccination — Kentucky, May–June 2021. MMWR Morb Mortal Wkly Rep. 2021;70(32):1081–1083. doi:10.15585/mmwr.mm7032e1

4. Hansen CH, Michlmayr D, Gubbels SM, Mølbak K, Ethelberg S. Assessment of protection against reinfection with SARS-CoV-2 among 4 million PCR-tested individuals in Denmark in 2020: a population-level observational study. The Lancet. 2021;397(10280):1204–1212. doi:10.1016/S0140-6736(21)00575-4

5. Altarawneh HN, Chemaitelly H, Hasan MR, et al. Protection against the Omicron Variant from Previous SARS-CoV-2 Infection. N Engl J Med. 2022;386(13):1288–1290. doi:10.1056/NEJMc2200133

6. Michlmayr D, Hansen CH, Gubbels SM, et al. Observed Protection Against SARS-CoV-2 Reinfection Following a Primary Infection: A Danish Cohort Study Using Two Years of Nationwide PCR-Test Data. SSRN Journal. Published online 2022. doi:10.2139/ssrn.4054807

7. Schmidt M, Schmidt SAJ, Adelborg K, et al. The Danish health care system and epidemiological research: from health care contacts to database records. Clin Epidemiol. 2019;11:563–591. doi:10.2147/CLEP.S179083

8. Danish Health Authority. Vaccination calendar. Danish Health Authority. Published January 8, 2021. Accessed February 23, 2022. https://www.sst.dk/-/media/English/Publications/2021/Corona/Vaccination/Target_groups_overview-_A4_colour_Engish.ashx?la=en&hash=DB051965FADA4DD4816755F5DE76210F8C23EC98

9. Schmidt M, Pedersen L, Sørensen HT. The Danish Civil Registration System as a tool in epidemiology. Eur J Epidemiol. 2014;29(8):541–549. doi:10.1007/s10654-014-9930-3

10. Schønning K, Dessau RB, Jensen TG, et al. Electronic reporting of diagnostic laboratory test results from all healthcare sectors is a cornerstone of national preparedness and control of COVID-19 in Denmark. APMIS. 2021;129(7):438–451. doi:10.1111/apm.13140

11. Sundhedsog Ældreministeriet. Danish Act no. 1615 on access to and registration of pharmaceutical and vaccination information of 18 December 2018 [Bekendtgørelse om adgang til og registrering m.v. af lægemiddelog vaccinationsoplysninger].; 2018:7. Accessed May 17, 2022. https://www.retsinformation.dk/api/pdf/205925

12. Schmidt M, Schmidt SAJ, Sandegaard JL, Ehrenstein V, Pedersen L, Sørensen HT. The Danish National Patient Registry: a review of content, data quality, and research potential. Clin Epidemiol. 2015;7:449–490. doi:10.2147/CLEP.S91125

13. Fernandez-Montero A, Argemi J, Rodríguez JA, Ariño AH, Moreno-Galarraga L. Validation of a rapid antigen test as a screening tool for SARS-CoV-2 infection in asymptomatic populations. Sensitivity, specificity and predictive values. EClinicalMedicine. 2021;37:100954. doi:10.1016/j.eclinm.2021.100954

14. Gram MA, Emborg HD, Schelde AB, et al. Vaccine effectiveness against SARS-CoV-2 infection and COVID-19-related hospitalization with the Alpha, Delta and Omicron SARS-CoV-2 variants: a nationwide Danish cohort study. medRxiv. Published online January 1, 2022:2022.04.20.22274061. doi:10.1101/2022.04.20.22274061

15. von Elm E, Altman DG, Egger M, et al. The Strengthening the Reporting of Observational Studies in Epidemiology (STROBE) statement: guidelines for reporting observational studies. Epidemiology. 2007;18(6):800–804. doi:10.1097/EDE.0b013e3181577654

16. Bartleson JM, Radenkovic D, Covarrubias AJ, Furman D, Winer DA, Verdin E. SARS-CoV-2, COVID-19 and the aging immune system. Nat Aging. 2021;1(9):769–782. doi:10.1038/s43587-021-00114-7

17. Collier DA, Ferreira IATM, Kotagiri P, et al. Age-related immune response heterogeneity to SARS-CoV-2 vaccine BNT162b2. Nature. 2021;596(7872):417–422. doi:10.1038/s41586-021-03739-1

18. Müller L, Andrée M, Moskorz W, et al. Age-dependent Immune Response to the Biontech/Pfizer BNT162b2 Coronavirus Disease 2019 Vaccination. Clinical Infectious Diseases. 2021;73(11):2065–2072. doi:10.1093/cid/ciab381

19. Nordström P, Ballin M, Nordström A. Risk of SARS-CoV-2 reinfection and COVID-19 hospitalisation in individuals with natural and hybrid immunity: a retrospective, total population cohort study in Sweden. The Lancet Infectious Diseases. Published online April 2022:S1473309922001438. doi:10.1016/S1473-3099(22)00143-8

20. Altarawneh HN, Chemaitelly H, Ayoub H, et al. Effect of prior infection, vaccination, and hybrid immunity against symptomatic BA.1 and BA.2 Omicron infections and severe COVID-19 in Qatar. medRxiv. Published online March 22, 2022:2022.03.22.22272745. doi:doi.org/10.1101/2022.03.22.22272745

21. Plumb ID, Feldstein LR, Barkley E, et al. Effectiveness of COVID-19 mRNA Vaccination in Preventing COVID-19–Associated Hospitalization Among Adults with Previous SARS-CoV-2 Infection — United States, June 2021–February 2022. MMWR Morb Mortal Wkly Rep. 2022;71(15):549–555. doi:10.15585/mmwr.mm7115e2

22. Ritchie H, Mathieu E, Rodés-Guirao L, et al. Coronavirus Pandemic (COVID-19). Our World in Data. Published online March 5, 2020. Accessed May 12, 2022. https://ourworldindata.org/coronavirus-testing

